# Current prices versus minimum costs of production for CFTR modulators

**DOI:** 10.1101/2022.01.29.22270088

**Authors:** Jonathan Guo, Junzheng Wang, Jingchun Zhang, Joseph Fortunak, Andrew Hill

## Abstract

**Background:** While the clinical benefits of CFTR modulators are clear, their high prices render them inaccessible outside of a select few countries. Despite this, there is currently limited evidence regarding access to these transformative therapies. Therefore, this study aims to estimate the minimum costs of production of CFTR modulators, assuming robust generic competition, and to compare them with current list prices to evaluate the feasibility of increased global access to treatment.

**Methods:** Minimum costs of production for CFTR modulators were estimated via an algorithm validated in previous literature and identification of cost-limiting key starting materials from published routes of chemical synthesis. This algorithm utilised per kilogram active pharmaceutical ingredient costs obtained from global import/export data. Estimated production costs were compared with published list prices in a range of countries.

**Results:** Costs of production for tezacaftor/ivacaftor/elexacaftor are estimated at $5676 [$4811-6539] per year, over 90% lower than the US list price. Analysis of chemical structure and published synthetic pathways for tezacaftor/ivacaftor/elexacaftor revealed relatively straightforward routes of synthesis related to currently available products. Total cost of triple therapy for all eligible diagnosed CF patients would be $489 million per year. Comparatively, the annual cost at US list price would be $31.2 billion.

**Conclusion:** Tezacaftor/ivacaftor/elexacaftor could be produced via generic companies for a fraction of the list price. The current pricing model restricts access to the best available therapy in LMICs, thereby exacerbating existing international inequalities in CF care. Urgent action is needed to increase availability of triple combination treatment worldwide.

**Funding:** None

## Introduction

In recent years, management for cystic fibrosis (CF) has advanced drastically, culminating in the development of CFTR modulators able to target the root cause of the condition. Four such drugs currently are licensed and sold by Vertex Pharmaceuticals: Kalydeco (ivacaftor), Orkambi (ivacaftor/lumacaftor), Symdeko (tezacaftor/ivacaftor) and Trikafta (tezacaftor/ivacaftor/elexacaftor). The most recent of these – the triple combination tezacaftor/ivacaftor/elexacaftor – is suitable for the largest proportion of mutation profiles (an estimated 90% of all CF patients)[1] and shows significantly improved outcomes when compared with both placebo and previous generation therapies.[2] These drugs currently represent the best available treatment for CF, and an opportunity to improve quality and length of life for almost all patients, yet US list prices for CFTR modulators are over $250,000 per year.[3] This pricing strategy renders the medicines out of reach for patients unless reimbursed by government or health system authorities. Even still, CFTR modulators pose resource allocation dilemmas to even the most robust and well-funded of health systems. This was seen most recently in the negotiations between Vertex and NHS England regarding prices for ivacaftor/lumacaftor which lasted four years and culminated in the destruction of 7,880 packs of expired drug.[4] The protracted negotiations caused by the pricing structure of CFTR modulators occur at the cost of patients, who remain in limbo unable to obtain life-changing treatments. As a result of this, some patients have even resorted to setting up “buyers clubs” to bypass both Vertex and regulatory bodies.[5]

The benefits of CFTR modulator therapy are so profound that such delays in access cause very tangible impacts on patient outcomes. For example, recent research has indicated universal introduction of tezacaftor/ivacaftor/elexacaftor in 2021 would reduce the number of people living with severe lung disease by 60% and deaths by 15% by 2030, with these benefits halved if introduction was delayed to 2025.[6]

Although much of the current documented CF burden lies in high-income countries (HICs), recent research suggests a high level of underreporting of CF in low- and middle-income countries (LMICs), due to ascertainment bias and lack of patient registries.[1,7] While such a pricing model delays access to treatment in HICs, it entirely disregards patients and health systems in LMICs where poorer outcomes are already experienced,[1] thereby exacerbating existing international inequalities in CF care. Currently CFTR modulators are almost exclusively available in the world’s richest countries – as of June 2021, tezacaftor/ivacaftor/elexacaftor was only reimbursed in 16 countries worldwide. Despite this, little evidence currently exists on patient access to CFTR modulators.

Similar challenges in treatment access have been overcome for other conditions. In the case of triple combination antiretroviral (ARV) therapy used for the treatment of human immunodeficiency virus, as well as direct-acting antivirals (DAAs) for treatment of hepatitis C, similarly prohibitive drug prices were reduced by over 90%.[8] This was achieved via generic competition introduced through use of voluntary licensing, with subsequent increased volume demand and production efficiency,[8,9] and allowed millions in resource-limited settings access to lifesaving drugs.

Such price decreases were deemed ambitious and infeasible at the time and yet have resulted in significantly improved treatment uptake and patient outcomes.[10] As such, using the precedents set by these pharmaceutical agents, this study aims to provide novel estimates of the minimum costs of production of CFTR modulators, assuming robust generic competition, and to compare them with current list prices to evaluate the feasibility of increased global access to treatment.

## Methods

### Minimum costs of production

Methodologies utilising prices of active pharmaceutical ingredient (API) have been described in previous research to reliably estimate minimum costs of production for standard oral formulations.[9,11,12] Since all CFTR modulator therapies on the market are taken PO, an algorithm adapted from Hill et al. was used to estimate costs of production. All four CFTR modulators with FDA and EMA approval were included within this study, though our analysis focused on ivacaftor/lumacaftor, tezacaftor/ivacaftor and tezacaftor/ivacaftor/elexacaftor due to their increased efficacy and patient eligibility when compared with ivacaftor monotherapy.[1]

Generic production in India (or China) was assumed as these are the world’s leading producers of both generic medications and API.[13] The import/export databases Panjiva and SINOIMEX, alongside PharmaCompass were used to analyse all available cost data for shipments of relevant API from 1^st^ January 2016 to July 2021. Shipments containing less than 1kg of API or API in combination with another product were excluded to avoid analysis of shipments of completed drug or samples not intended for large-scale production. Duplicates were removed and shipments with a price per kilogram outside of the 15^th^ and 85^th^ percentiles excluded to reduce the effect of outliers on our estimates. After completion of our analysis, we included the data outside the 15^th^ and 85^th^ percentiles to assure that these exclusions did not bias results due to any unusual circumstances.

Weighted-mean price per kilogram of API was calculated and combined with dosage information extracted from the British National Formulary (BNF). This was used to calculate API costs for a one-year course of treatment, including an assumed 5% API loss during production. Total production cost was calculated by incorporating costs of excipients (substances required for formulation such as stabilisers, binding agents, disintegration/dissolution enhancers, and coatings) and formulation into a tablet for oral consumption. Previous research has estimated average pharmaceutical excipient cost to be $2.63 per kilogram of finished pharmaceutical product (FPP).[14] Similarly, per-unit formulation cost was estimated at $0.01 per tablet.[11,15] Finally, a profit margin of 10% was added, as well as an average Indian tax of 27% to calculate FPP price.[16].

No API data was available for elexacaftor, likely owing to its novelty. Consequently, for this API estimates of per kilogram costs were made via published routes of chemical synthesis extracted from patent information.[17] Similar analyses were performed based upon chemical structure and synthetic routes for other APIs to identify cost-limiting key starting materials (KSMs) and ensure estimated costs of production were reasonable.

### Current list prices

National pricing databases in a variety of countries were searched in August 2021 to provide a snapshot of current publicly available information on list prices, calculated for one-year treatment courses. These were then compared with our estimated minimum costs of production. As it currently represents the largest market for CFTR modulators, for the US, both commercially available price to the public, and costs to the publicly funded veteran’s affairs system were listed. Costs from the UK’s CF Buyers’ Club, which enables patients to buy low-cost generic CFTR medications produced in Argentina, are also quoted for completeness. The full list of data sources by country are detailed in Appendix 1.

Where official databases were unavailable, online pharmacy sites were used as an alternative. Where several prices were available in the same database, the lowest was selected. Differences in medicine composition were accounted for between regulatory jurisdictions, with standardisation of pricing based on the dosing regimen found in the BNF.

## Results

### Minimum costs of production

Weighted-mean API prices for ivacaftor, lumacaftor and tezacaftor were found to be $13,196/kg, $19,673/kg and $51,500/kg respectively. Overall, this study identified shipment volumes of 186.93kg for ivacaftor in total, with 125.96kg being in the latest 12-month period. This was an increase of 314.48% compared to the average annual volume of 30.39kg since 2016. In terms of lumacaftor, total shipment volumes of 37.76kg were identified, with 16.71kg in the last 12-months and the average annual volume was found to be 5.21kg since 2016. It should be noted that after data cleaning only one shipment result was available for tezacaftor which influences the reliability of this estimate.

Ivacaftor API was shipped from India to the US in 2015 (40 kg) at an average cost of $30,889/kg.[18] This likely represents shipments of API to Vertex in the earlier stages of commercialization. Eight companies (seven in India) have Drug Master Files (DMFs) on this drug with the USFDA.[19] Seven of these companies are Indian; six of them appear capable of making the API while one company (Vindhya) seems to be supplying key starting materials for API production. Eight additional companies advertise their ability to supply the API (without a US or European registration) on the website PharmaCompass. The largest importer of API is Argentina, at an average cost of $12,700/kg.[20]

There is only a single Drug Master File listed for lumacaftor with two API suppliers listed on PharmaCompass. Tezacaftor also has only one listed DMF with the FDA, with three companies listed on PharmaCompass as suppliers of the API. No reliable pricing data is available for these APIs in PharmaCompass.

Analysis of chemical structure and published synthetic pathways for elexacaftor revealed relatively straightforward routes of synthesis. The API is made, however, by putting together multiple modular pieces (starting materials) that are fairly expensive because they are not in high demand for other uses in the chemical industries. Consequently, API cost was estimated to be $20,000/kg. A margin of error of ±$10,000/kg was included to allow for uncertainty and consequently produce additional values for the final drug price of tezacaftor/ivacaftor/elexacaftor. All schema and chemical structures for relevant APIs and KSMs can be found in Appendices 2-6.

### Orkambi – ivacaftor/lumacaftor

The treatment regimen for ivacaftor/lumacaftor is one 250/400mg tablet taken twice daily. Thus, for a one-year treatment course of ivacaftor/lumacaftor total API cost was $8153 (Figure 1a). With incorporation of production costs, the total estimated cost of production for one year of treatment was $9659. Our model renders ivacaftor/lumacaftor the most expensive of the CFTR modulators to produce. This is likely due to the relatively high daily dosage of API contained within the treatment.

**Figure 1a.**
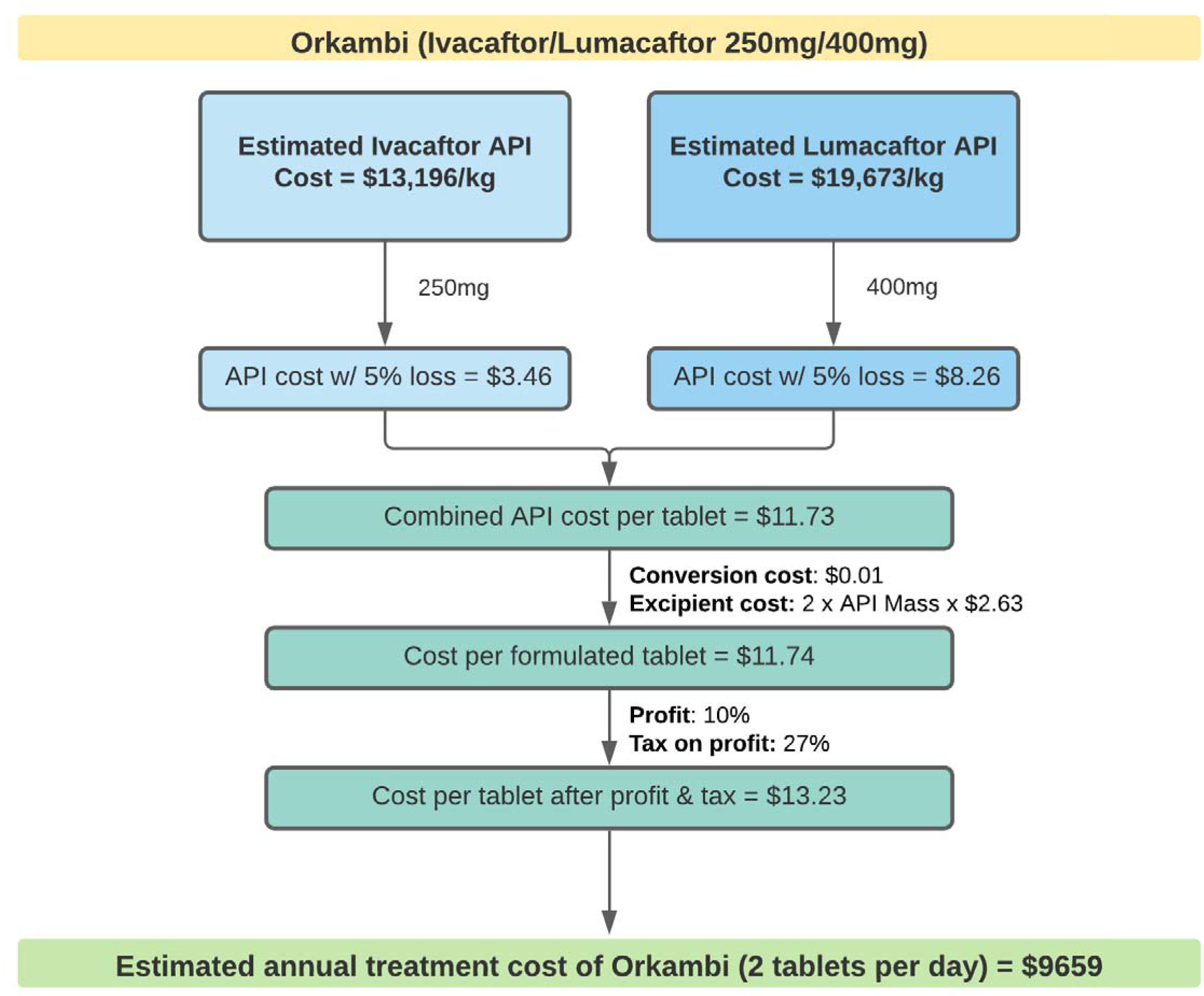
Flowchart showing minimum cost estimations for Orkambi

The synthesis of the API ivacaftor is relatively straightforward. In addition to the originator (Vertex), other research groups have published synthetic routes for this molecule.[21,22] These approaches have been reviewed.[17] Two key intermediates are used to synthesize ivacaftor. In any of the several published syntheses, intermediate 9 or 10 is a key starting material (Appendix 3, Scheme 1).

The most straightforward way of synthesizing ivacaftor is by peptide coupling of quinolone-3-carboxylic acid 4 with 9 or 10 (Scheme 1). There are several ways of preparing 4 from inexpensive and readily available starting materials using known chemistry. This approach suggests that 4 and 10 can be sourced as KSMs, and potentially minimizes the number of steps in the API synthesis that require processing under Good Manufacturing Practice (GMP). Processing under GMP is more expensive than making compounds for the fine chemicals industry due to the enhanced level of testing, validation, and documentation required. Alternative syntheses have been published that avoid patented chemistry or intermediates (Scheme 2). Each of these routes has the potential to provide the API on reasonable (metric ton) scale at much less than the current pricing shown in import/export data.

Intermediates 42, 43, and 51 are the KSMs for synthesis of the API lumacaftor (Appendix 4). Schemes 3, 4, and 5 represent a favourable route for combining these intermediates to synthesize the API. This is a much higher-yielding and more efficient route than the synthesis originally disclosed by Vertex for early clinical development (Scheme 6).

### Symdeko – tezacaftor/ivacaftor

Daily treatment with tezacaftor/ivacaftor entails one 100/150mg tablet followed by one 150mg tablet of ivacaftor. Therefore, estimated total API cost for one year of treatment was $3325, with a final annual treatment cost of $3943 after accounting for extra costs of production (Figure 1b).

**Figure 1b.**
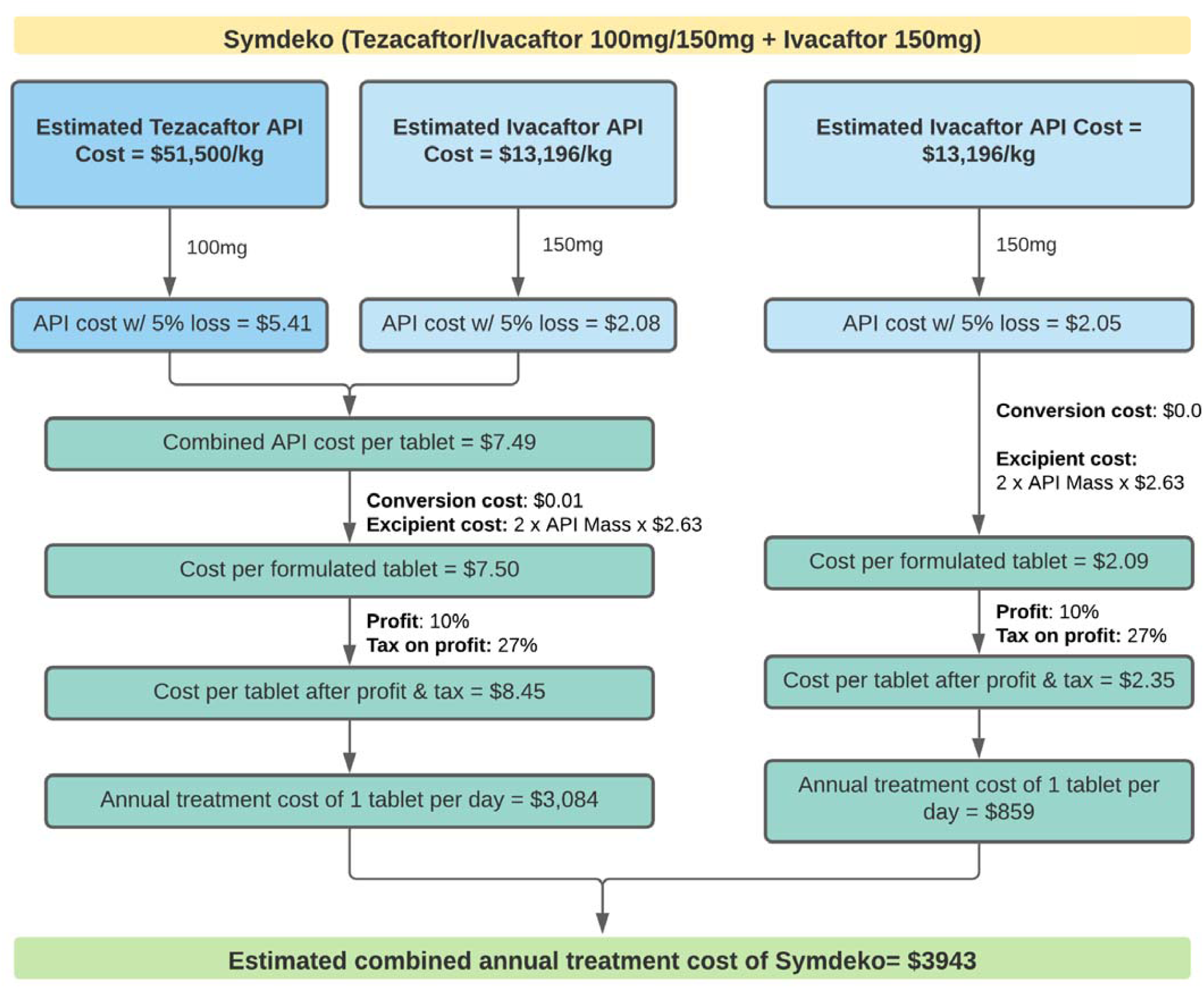
Flowchart showing minimum cost estimations for Symdeko

Compounds 97, 98 (Appendix 5, Schemes 8 and 9), 49, and R-(benzyl)glycidol are KSMs for the synthesis of tezacaftor API. Intermediate 49 is in effect a common intermediate for the synthesis of lumacaftor and is prepared from 43.

### Trikafta – tezacaftor/ivacaftor/elexacaftor

Daily treatment with tezacaftor/ivacaftor/elexacaftor consists of two 50/75/100mg tablets followed by one 150mg tablet of ivacaftor. As such, estimated total API cost for one year of treatment was $4785. After accounting for production costs, the final estimated annual cost of treatment was $5676 (Figure 1c). With the margin of error of ±$10,000/kg API, the highest estimated cost was $6539, with the lowest at $4811.

**Figure 1c.**
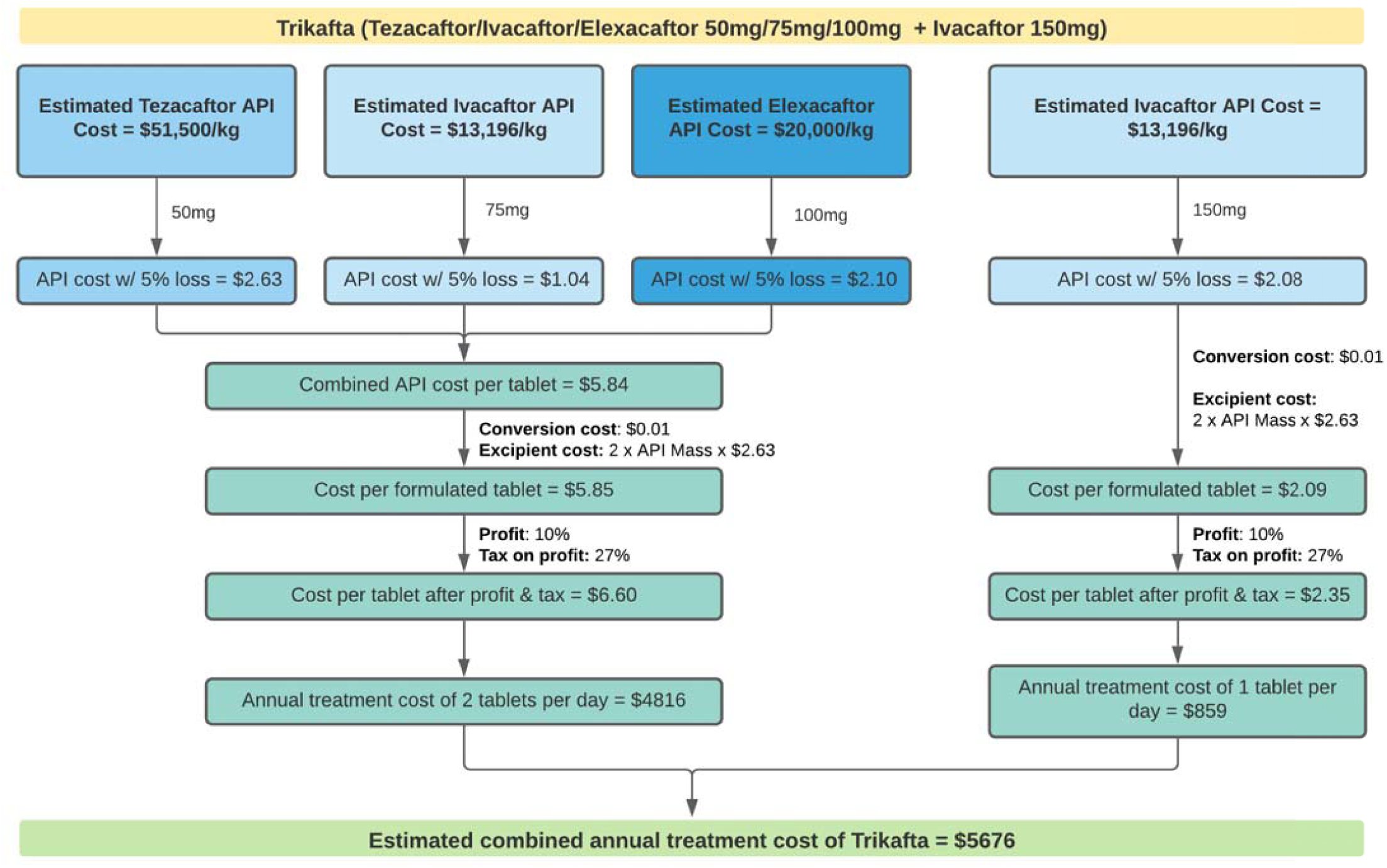
Flowchart showing minimum cost estimations for Trikafta

The KSMs for synthesizing elexacaftor API are respectively, compounds 97, 109, 116, 123, and 125 (Appendix 6). No alternative synthetic routes to those disclosed by Vertex have so far been published.

### Current list prices for CFTR modulators

Our results are summarised in Table 1 and Figure 2a-c. Overall, available data is limited to countries where CFTR medications have been approved, which tended to be in western HICs with a relatively high burden of diagnosed disease, and where healthcare systems have the resources available to afford such drugs.

**Table 1.** Summary of list prices vs estimated production costs for CFTR combination therapies. Costs rounded to nearest $100. OD = once daily, BD = twice daily.

| Drug, Duration and Dose | Highest List Price<br>(per annum) | Lowest List Price<br>(per annum) | Estimated<br>Production Cost<br>(per annum) |
| --- | --- | --- | --- |
| **Orkambi**<br>**PO ivacaftor/lumacaftor**<br>250/400mg BD | \$284,800<br>US (Commercial) | \$79,900<br>Argentina | \$9,700 |
| | | \$15,000<br>UK CF Buyers' Club | |
| **Symdeko**<br>**PO tezacaftor/ivacaftor**<br>100/150mg OD + ivacaftor 150mg OD | \$305,000<br>US (Commercial) | \$94,300<br>Argentina | \$3,900 |
| | | \$15,000<br>UK CF Buyers' Club | |
| **Trikafta**<br>**PO tezacaftor/ivacaftor/elexacaftor**<br>100/150/200mg OD + ivacaftor 150mg OD | \$325,300<br>US (Commercial) | \$102,300<br>Argentina | \$5,700 |

**Figure 2a.**
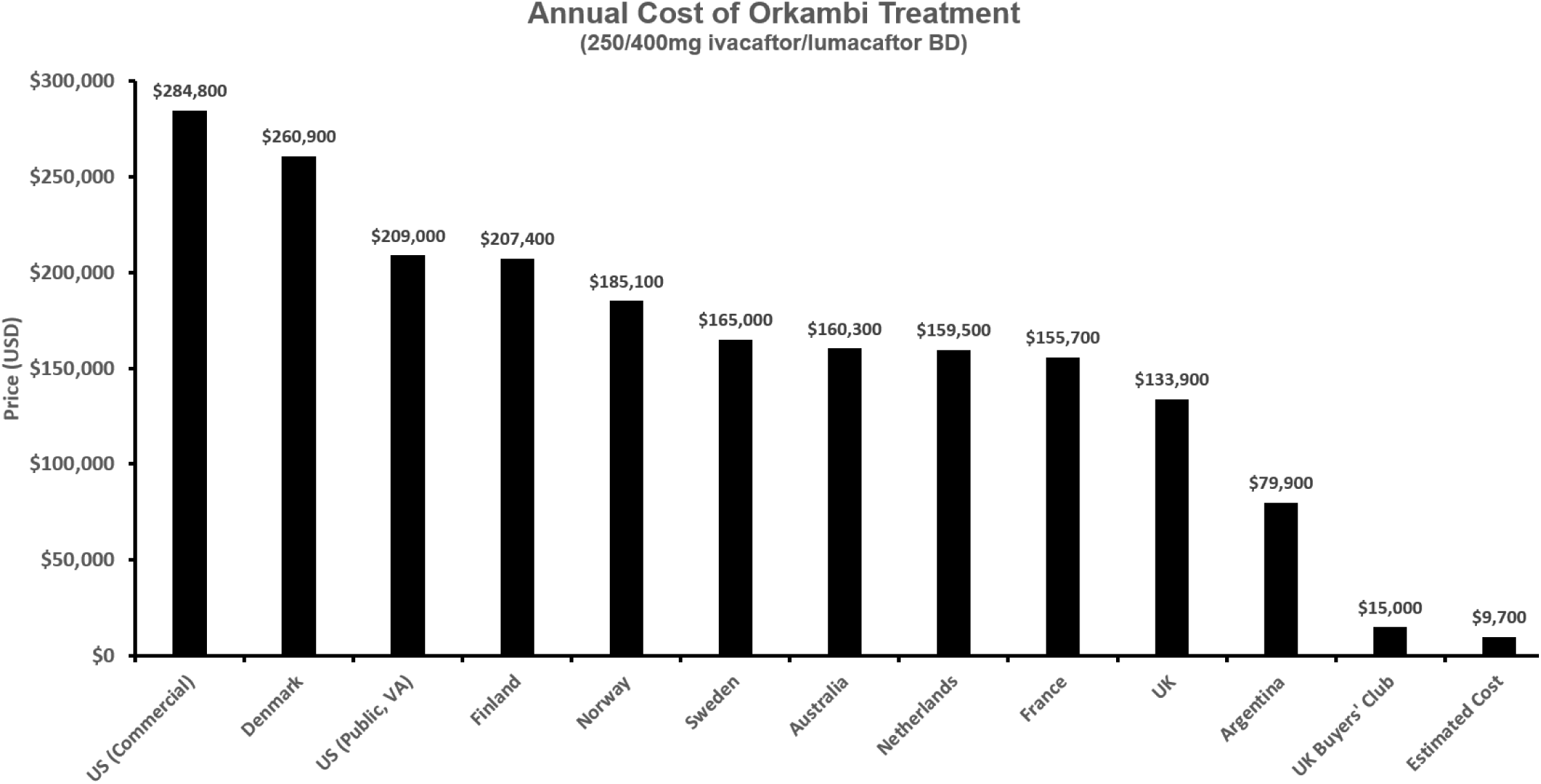
Graph of national treatment course prices of Orkambi compared to generic estimate

**Figure 2b.**
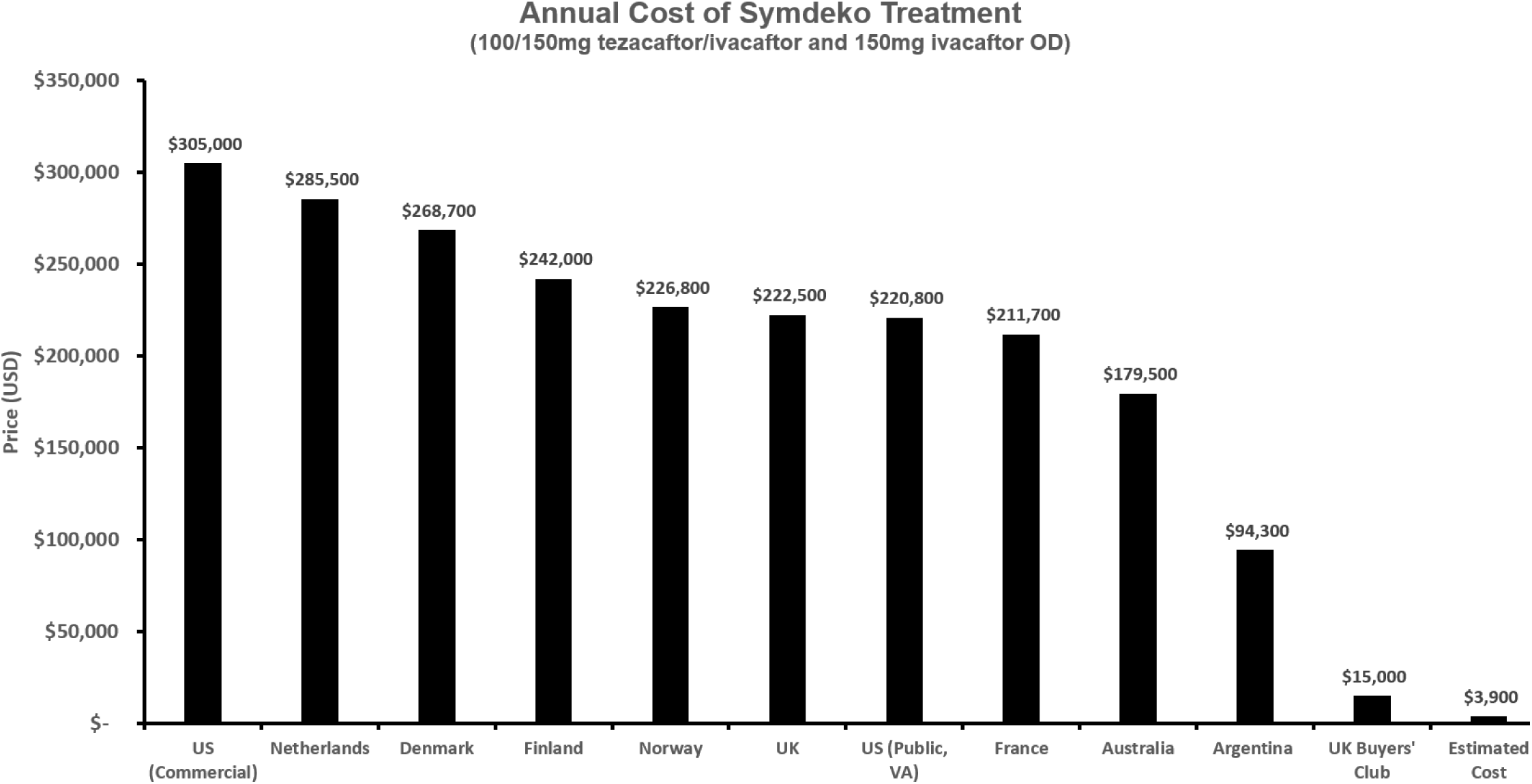
Graph of national treatment course prices of Symdeko compared to generic estimate

**Figure 2c.**
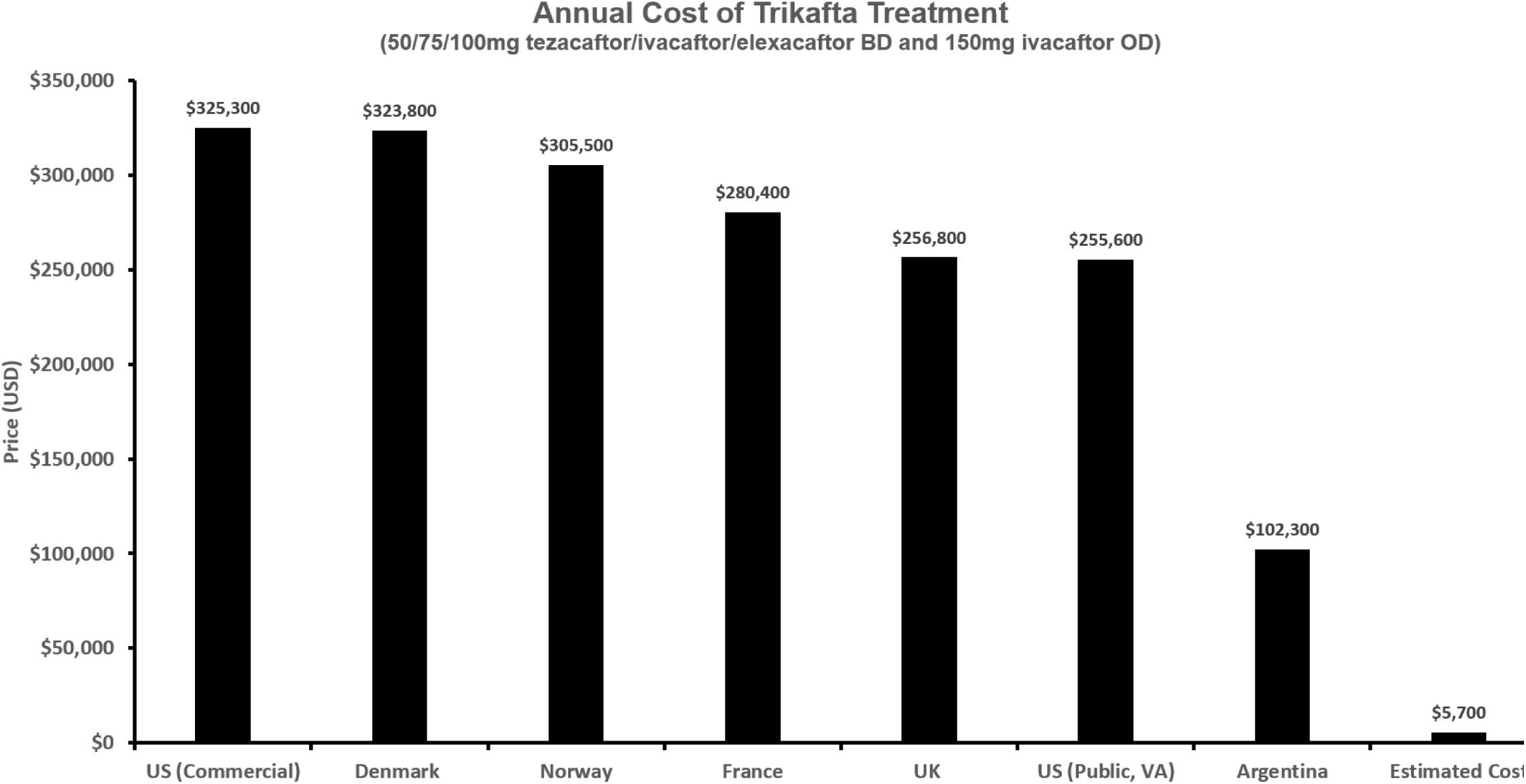
Graph of national treatment course prices of Trikafta compared to generic estimate

For each combination treatment the highest international price found was via commercial US pharmacy, and the lowest in Argentina. This is likely because in Argentina CFTR modulators are currently not under patent protection, and as such generic versions are available. In all cases commercial prices around the world were many times higher than the costs estimated by our model.

## Discussion

This novel analysis suggests that costs of production for CFTR modulators could be significantly lower than current list prices. Specifically, for the current best available CF treatment (tezacaftor/ivacaftor/elexacaftor), the lowest originator cost for one year of treatment was $255,600, while the highest production cost estimated by this analysis was $6539, representing a decrease of over 90%.

Generic versions of all CFTR modulator therapies are currently produced in Argentina, where patent restrictions currently do not apply. Under the Trade-Related Aspects of Intellectual Property Rights agreement commercial export of generic drugs to a country where such products remain under patent protection is not permitted.[23] As such in this rather unique case the market for generic CFTR modulators is mostly limited to domestic Argentinian CF patients, and there is an absence of the competition usually required to drive down drug prices.[8] Yet even with a limited volume demand and the absence of competition, these generics formed the lowest list price for every modulator combination and can be purchased for prices of ∼$15,000,[5,24] indicating the commercial viability of lower cost CFTR modulators and the reliability of the minimum prices reached by this analysis. The methodologies used have also been previously validated to reliably estimate minimum costs of production (assuming robust generic competition) for a wide variety of other drugs such as those on the WHO essential medicines list, insulin analogues and DAAs.[9,11,12]

Several limitations should be noted. Firstly, our model assumes generic production in specific geographical locations with robust mechanisms of competition. Accordingly, it does not account for potentially required investment in new facilities, regulatory approval or costs of research and development of products.

In previous analyses utilising similar methodologies higher volumes of API were identified. This is likely due to both the novelty and limited manufacture of this class of drugs while they remain under patent. While the lower volumes within our analysis limit the accuracy of our estimates, larger scale production of both API and FPP generally sees prices fall rather than rise due to increased volume demand, process optimisation and production efficiencies.[9]

In addition, actual prices paid for CFTR modulators are likely lower than “list price” following negotiations with payors. While such data is not publicly available, previous research has found discounts for pharmaceuticals tend to range from 20-29%,[25] which still places the final prices identified by our survey significantly higher than those estimated in our analysis. As such even if, for example, a significantly higher profit margin were incorporated into the model to incentivise market entry for generic competitors, the final cost would remain a fraction of contemporary prices.

Cost-effectiveness analyses by both governmental and independent bodies have concluded that the benefits of CFTR modulators do not warrant their prices.[3,26] While pricing of orphan drugs does pose a challenge in that costs of development need to be recouped from a comparatively smaller population, even with the current limited rollout of tezacaftor/ivacaftor/elexacaftor, it is estimated to be one of the most valuable orphan drugs currently available,[27] with quarterly revenues reaching $1.5 billion.[28] It therefore appears prices are not determined via production expenses or efficacy, but rather what the market will bear.

At the time of writing, the total number of CF patients documented in publicly-reporting registries amounted to 95,835. Assuming this volume demand and a patient eligibility of 90%,[1] the annual cost of triple therapy for all eligible patients at the US list price would amount to $31.2 billion.[3] Even assuming negotiated discounts, such a cost represents an enormous barrier to equitable access to treatment, especially given that this estimate is mainly limited to HICs. Comparatively, at the calculated minimum costs of production this could be achieved for $489 million.

Patent protection for tezacaftor/ivacaftor/elexacaftor is expected to last until 2037.[3] Consequently, the current monopoly on modulator treatment is not set to change. It seems there is no transparent plan to increase access to these transformative therapies, as Vertex have largely not sought regulatory approval for modulator therapies outside of the Global North, despite the known disease burden of CF in many countries outside of this region.[1] As such unless steps are taken urgently to address current inequities, CFTR modulators will remain inaccessible to patients outside of the world’s richest countries, and disparities in outcomes will likely only grow larger.

Currently compassionate treatment programs are run by Vertex to provide treatments at lower cost,[29] however it is clear action is needed at a country, rather than individual level. With generic commercialisation of CFTR modulators already shown to be feasible in Argentina, one key mechanism used to increase access to ARVs and DAAs which could be applied to CFTR modulators is voluntary licensing. Aside from the policy’s previous success with other drugs, this strategy is preferable to such price reductions or donation programs as it generates a market for generic competition, thereby providing long-term, sustainable price decreases.[30]

## Conclusion

Country-level reimbursement of CFTR modulators falls far short of global disease burden. This is driven by their prohibitively high prices and has led to patients being neglected from advancements in care while perpetuating international disparities in CF outcomes. CFTR modulators could be produced for a fraction of current list prices. As such, urgent action to increase the availability of treatment is needed to prevent a widening of existing inequalities. One strategy based on previous success is originator-issued voluntary licenses.

## Data Availability

All data used in this study will be available from the corresponding author, upon reasonable request.

## Credit Author Statement

**Jonathan Guo:** Data curation, Formal analysis, Investigation, Methodology, Visualisation, Writing – Original Draft, Writing – Review & Editing. **Junzheng Wang:** Formal analysis, Visualization, Writing – Review & Editing. **Jingchun Zhang:** Data curation, Writing – Review & Editing. **Joseph Fortunak:** Data curation, Methodology, Formal analysis, Writing – Original Draft, Writing – Review & Editing. **Andrew Hill:** Conceptualization, Methodology, Supervision, Writing – Review & Editing.

## Ethical approval

N/A

## Acknowledgements

The authors would like to acknowledge the kind support of Rob Long, Gayle Pledger and Christina Walker and Susannah Wang.

