## Supplementary Appendix for "Current prices versus minimum costs of production for CFTR modulators"

**Appendix 1: List of national list price sources**

| **Source** | **Link** |
| --- | --- |
| **Argentina**  Kairos Argentina | <https://ar.kairosweb.com/> |
| **Australia**  Department of Health, Pharmaceutical Benefits Scheme | <http://www.pbs.gov.au/pbs/home> |
| **Denmark**  Danish Medicines Agency | [Medicinpriser.dk](http://medicinpriser.dk/) |
| **Finland**  Social Insurance Institution of Finland, Medicinal Products Database | <https://asiointi.kela.fi/laakekys_app/LaakekysApplication> |
| **France**  Ministry of Solidarity and Health, Database of Public Medicines | <http://base-donnees-publique.medicaments.gouv.fr/> |
| **Netherlands**  Netherlands Care Institute | <https://www.medicijnkosten.nl/> |
| **Norway**  Norwegian Medicines Agency, NoMA Medicine Database | <https://www.legemiddelsok.no/> |
| **Sweden**  Dental Care and Medicines Benefit Agency, Drug Database | https://www.tlv.se/beslut/sok-i-databasen.html |
| **United Kingdom**  British National Formulary (BNF) | <https://bnf.nice.org.uk/drug/> |
| **United Kingdom**  Cystic Fibrosis Buyer’s Club | <https://www.cfbuyersclub.org/> |
| **United States (Commercial)**  Drugs.com | <https://www.drugs.com/> |
| **United States (VA)**  Department of Veteran Affairs, Office of Procurement, Acquisition and Logistics (OPAL) | <https://www.va.gov/opal/nac/fss/pharmPrices.asp> |

**Appendix 2 – Table displaying chemical structures of CFTR modulator active ingredients and key starting materials required for synthesis**

| CFTR Modulator Active Ingredient | Identified Key Starting Materials |
| --- | --- |
| **Ivacaftor**  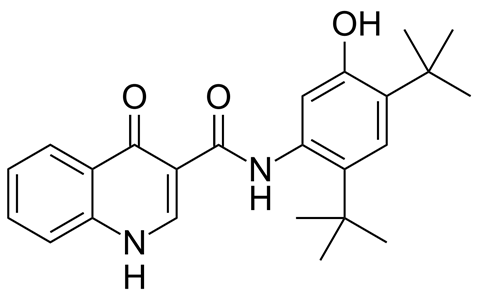 | 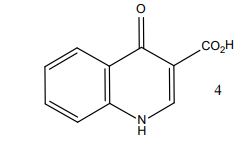  +  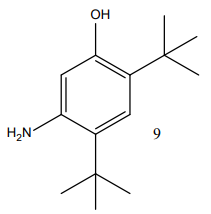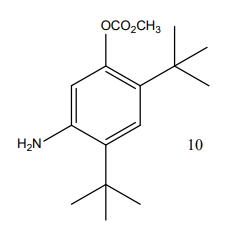  + |
| **Lumacaftor**  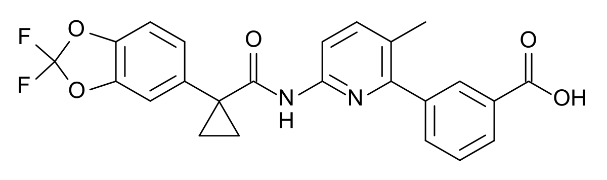 | 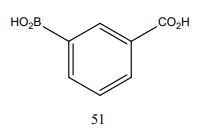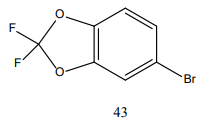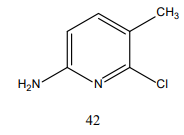  +  + |
| **Tezacaftor**  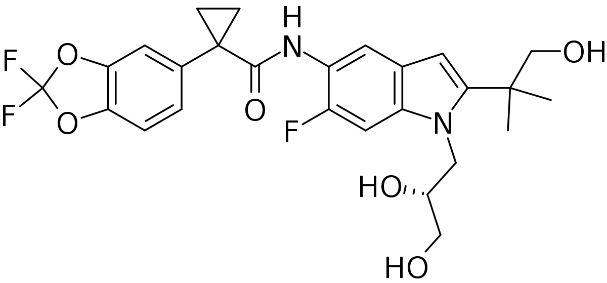 | 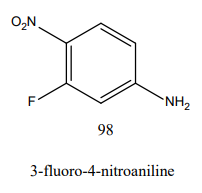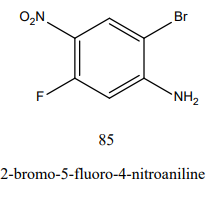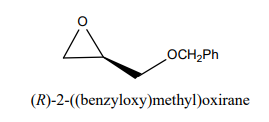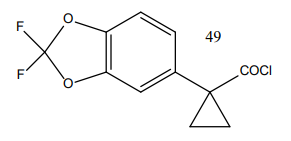  +  +  +  +  +  + |
| **Elexacaftor**  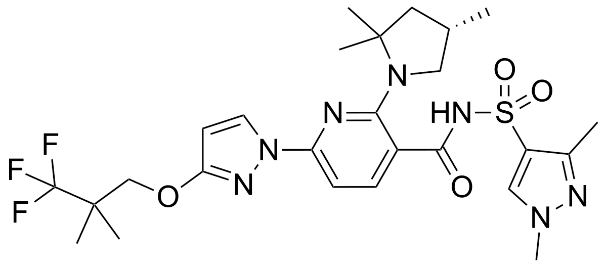 | 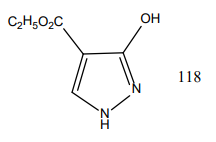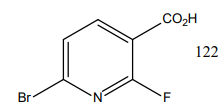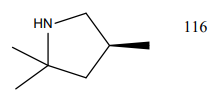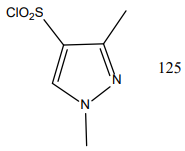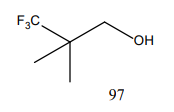  +  +  +  +  +  +  +  + |

**Appendix 3 – Relevant schema and key starting materials for synthesis of ivacaftor**

Scheme 1. General synthetic approach to ivacaftor

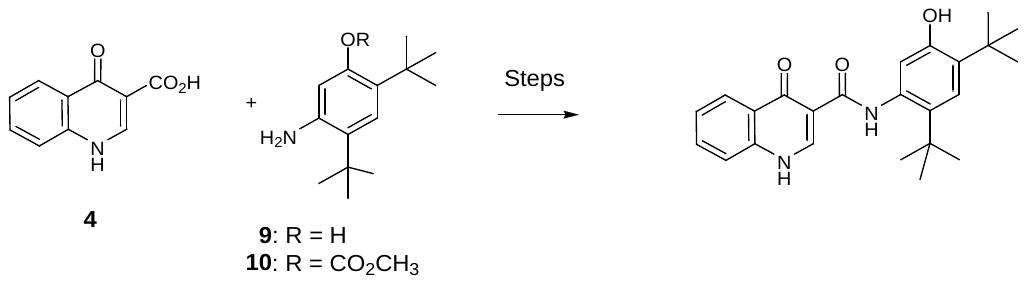

Scheme 2. Alternative synthesis of ivacaftor

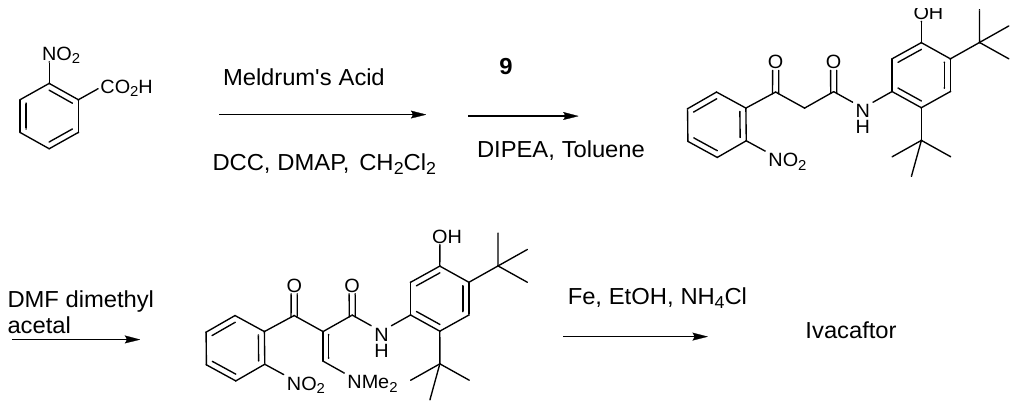

**Appendix 4 – Relevant schema and key starting materials for synthesis of lumacaftor**

Scheme 3. Synthesis of KSM 42 for lumacaftor

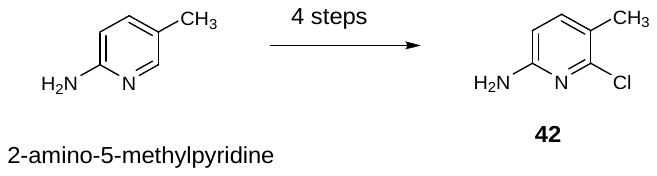

Scheme 4. Synthesis of intermediate 49 from KSM 43 for lumacaftor

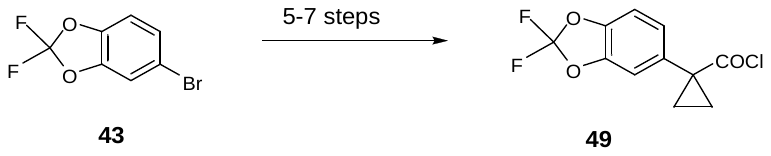

Scheme 5. Second-generation synthesis of lumacaftor.

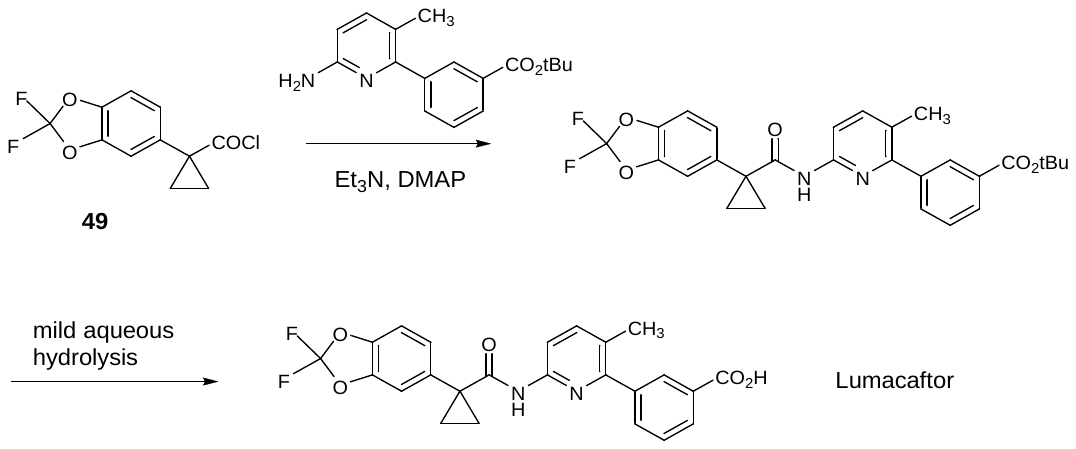

Scheme 6. First-Generation synthesis of lumacaftor

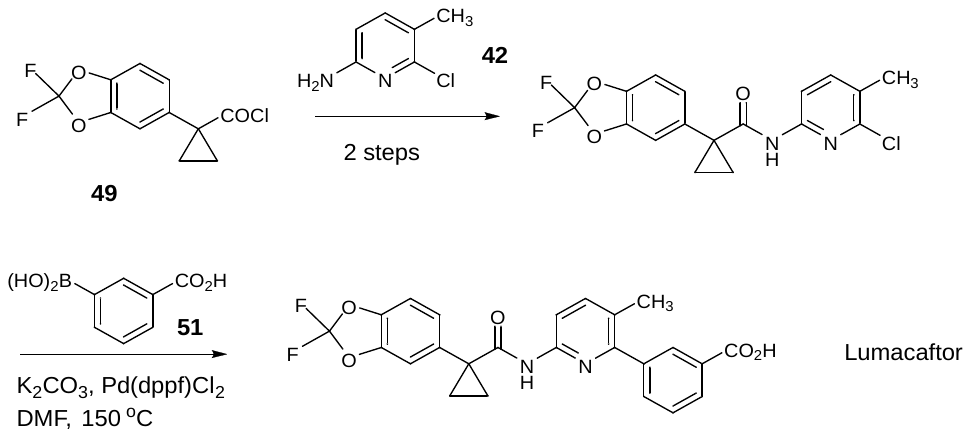

**Appendix 5 – Relevant schema and key starting materials for synthesis of tezacaftor**

Scheme 7. Synthesis of intermediate 84 for tezacaftor.

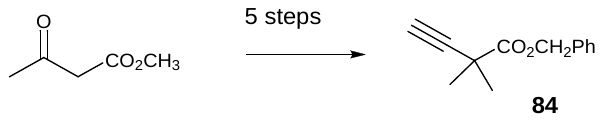

Scheme 8. Synthesis of intermediate 97 for tezacaftor.

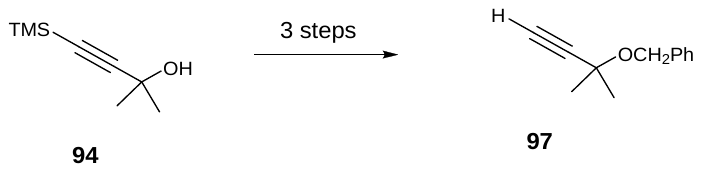

Scheme 9. Synthesis of intermediate 85 for tezacaftor.

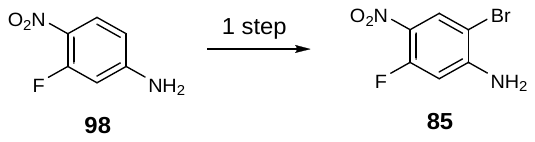

Figure 1. Key starting materials 88 and 89 for tezacaftor

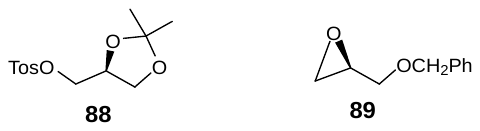

Scheme 10. First generation synthesis of tezacaftor.

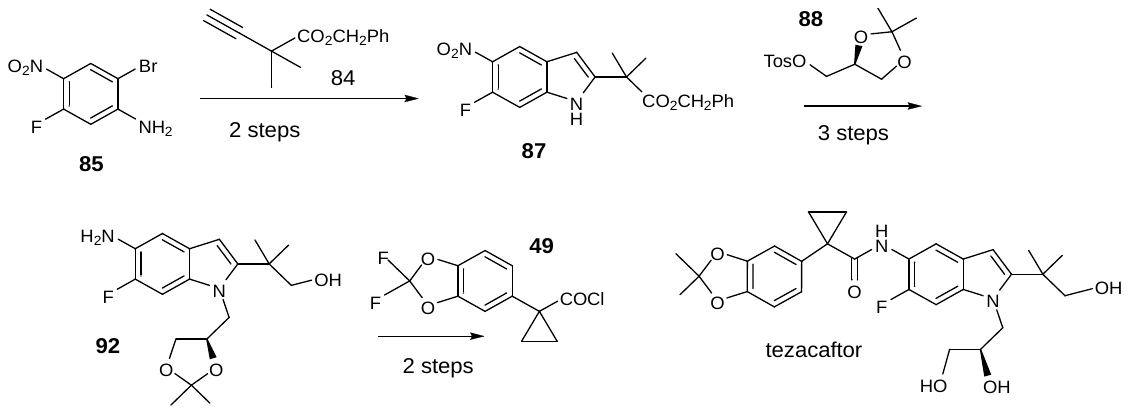

Scheme 11. Second generation synthesis of tezacaftor.

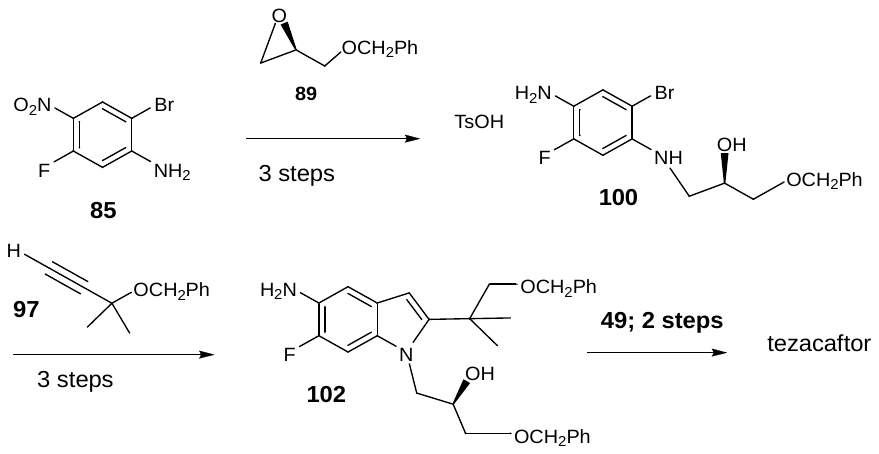

**Appendix 6 – Relevant schema and key starting materials for synthesis of elexacaftor**

Figure 2. Key starting materials for elexacaftor synthesis

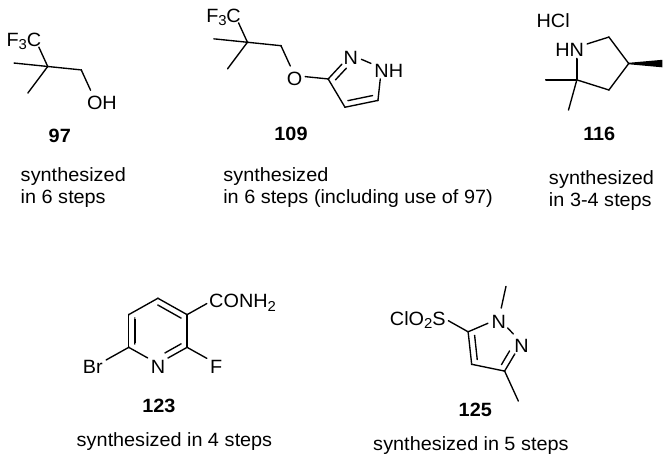

Scheme 12. Synthesis of elexacaftor

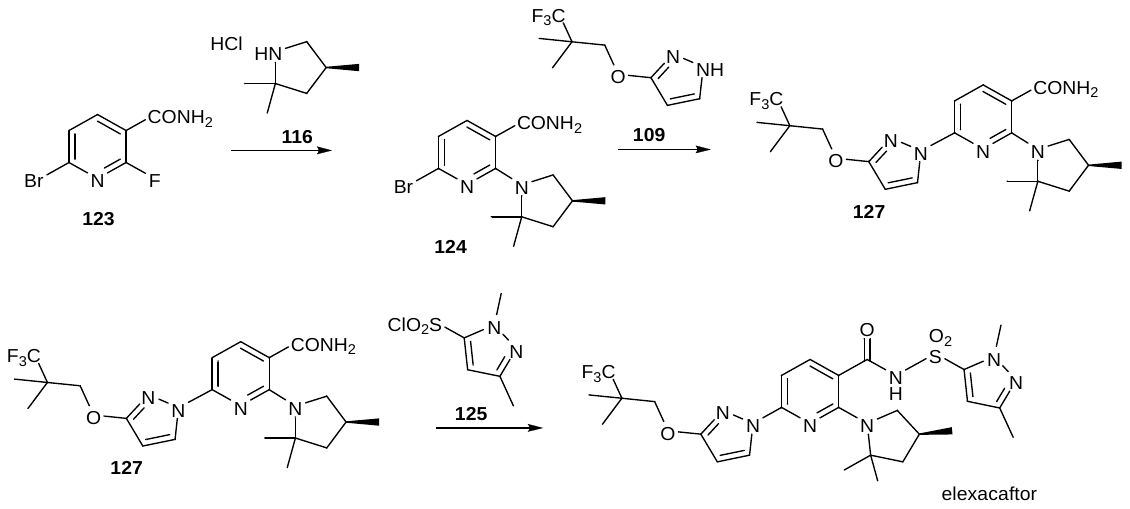
